# Determinants of SARS-CoV-2 Vaccine Engagement in Algeria: A Population-based Study with Systematic Review of Studies from Arab Countries of the MENA Region

**DOI:** 10.1101/2021.07.17.21260662

**Authors:** Salah Eddine Oussama Kacimi, Selma Nihel Klouche-Djedid, Omar Riffi, Hadj Ahmed Belaouni, Farah Yasmin, Fatma Asma Taouza, Yasmine Belakhdar, Saliha Chiboub Fellah, Amira Yasmine Benmelouka, Shoaib Ahmed, Mohammad Aloulou, Abdellah Bendelhoum, Hafida Merzouk, Sherief Ghozy, Mohammad Yasir Essar, Mohamed Amine Haireche

**Author notes:** Contributed equally (Salah Eddine Oussama Kacimi, Selma Nihel Klouche-Djedid).

## Abstract

**Background:** The Algerian COVID-19 vaccination campaign, which started by the end of January 2021, is marked by a slowly ascending curve despite the deployed resources. To tackle the issue, we assessed the levels and explored determinants of engagement towards the COVID-19 vaccine among the Algerian population.

**Methods:** A nationwide, online-based cross-sectional study was conducted between March 27 and April 30, 2021. A two-stage stratified snowball sampling method was used to include an equivalent number of participants from the four cardinal regions of the country. A vaccine engagement scale was developed, defining vaccine engagement as a multidimensional parameter (5 items) that combined self-stated acceptance and willingness with perceived safety and efficacy of the vaccine. An Engagement score was calculated and the median was used to define engagement versus nonengagement. Sociodemographic and clinical data, perceptions about COVID-19 and levels of adherence to preventive measures were analyzed as predictors for nonengagement.

**Results:** We included 1,019 participants, 54% were female and 64% were aged 18-29 years. Overall, there were low rates of self-declared acceptance (26%) and willingness (21%) to take the vaccine, as well as low levels of agreement regarding vaccine safety (21%) and efficacy (30%). Thus, vaccine engagement rate was estimated at 33.5%, and ranged between 29.6-38.5% depending on the region (p>0.05). Nonengagement was independently associated with female gender (OR=2.31, p<0.001), low adherence level to preventive measures (OR=6.93p<0.001), private sector jobs (OR=0.53, p=0.038), perceived COVID-19 severity (OR=0.66, p=0.014), and fear from contracting the disease (OR=0.56, p=0.018). Concern about vaccine side effects (72.0%) and exigence for more efficacy and safety studies (48.3%) were the most commonly reported barrier and enabler for vaccine acceptance respectively; whereas beliefs in the conspiracy theory were reported by 23.4%.

**Conclusions:** The very low rates of vaccine engagement among the Algerian population probably explain the slow ascension of the vaccination curve in the country. Vaccine awareness campaigns should be implemented to address the multiple misconceptions and enhance the levels of knowledge and perception both about the disease and the vaccine, by prioritizing target populations and engaging both healthcare workers and the general population.

## Introduction

Amid the ongoing COVID-19 pandemic and the lack of effective curative treatments, mass vaccination is perceived as the only effective strategy to control the pandemic and reduce its global impact on individuals and societies. Different types of COVID-19 vaccines have been developed so far, using different techniques including mRNA, adenovirus vector, adjuvanted protein or live-attenuated or inactivated virus vaccines. The current evidence supports the efficacy of majority of the commercialized and recommended vaccines in eliciting robust production of neutralizing antibodies in a short- and median-term, correlating with a significant reduction in the incidence of COVID-19 infection both in clinical trial and real life (1–4).

As of 14 July 2021, the number of vaccine doses that have been administered is estimated at more than 3.5 billion, covering approximately 26% of the world population. However, there is a great discrepancy in vaccination rates between the industrialized countries such as Canada, the United Kingdom and the European countries, and developing and low-income countries (5). On the other hand, the recent emergence and spread of novel viral variants, notably the Indian (B.1.617) and Brazilian (P.1) variants, compromise the forecasted transition, on the short run, to the pre-pandemic normal life (6–10). As a consequence, the resolution of the issue depends on a three-fold concern, including the success of the global mass immunization, the long-term efficacy of the vaccines, and the dreaded scenario of resistance of the emerging variants to the vaccine-induced immunity (11–13).

In addressing the determinants of success for this global strategy, people’s engagement to local vaccination campaigns constitutes a major determinant, beside the adherence to prevention policies and recommendations. Although the modern experience with mass vaccination proved to be effective in controlling and eradicating outbreaks such as Polio, Smallpox, and other diseases (14), vaccine hesitancy has long been identified as one of the major threats facing the global health (15–17). Due to several factors, the COVID-19 vaccine is subject to recurrent popular misconceptions and uncertainties, which constitutes further barriers to public adherence with the vaccination strategy (18). Such misconceptions are reported to be particularly prevalent in developing countries and conservative societies, associated with high rates of vaccine hesitancy (19).

In Algeria, the largest African country, the fight against the virus has gone through successive phases since the first confirmed case declared on February 25, 2020. Since the early phase of the pandemic, the Algerian government opted for broad travel cancellations combined with intermittent implementation of restrictive and semi-restrictive measures locally, in addition to the deployment of tremendous healthcare resources to treat the infected citizens (20–22). As of 21 May 2021, the country has recorded 126,434 confirmed cases and 3,405 deaths (23). The national vaccination campaign started by the end of January 2021 and the current local policy targets all vulnerable groups. However, the vaccination rate remains remarkably low, reaching only 2.5 million doses by 14 July 2021, which represents a coverage rate estimated at 5.8% of the population (5,24). This contrasts with the country’s success in past immunization programs that enabled eradication and control of several communicable and zoonotic disease, and which developed the country a healthy one in the region (25,26).

In an attempt to explain this low vaccination rate, the present study was designed to evaluate the levels of engagement among Algerians towards the COVID-19 vaccine and to analyze the associated sociodemographic factors. Additionally, it explored the associated misconceptions and eventual barriers and enablers of vaccine acceptance. Such data would assist the decisionmakers in implementing strategic amendments on the vaccination policy and the related communication approaches. We further conducted a systematic review on vaccine acceptance in the Arab countries of the Middle-East and North African (MENA) region.

## Methods

### I. Cross-sectional study

#### I.1. Design & Population

A nationwide online-based cross-sectional study was conducted among the general population of Algeria, between March 27 and April 30, 2021. It involved adult (aged 18 years and older) males and females of all regions, who were permanently residing inside the country during the study period. Individuals who had previously received COVID-19 vaccine were excluded as shown in the flowchart in **Figure 1**. The study was approved by the institutional review board of the University of Tlemcen [14/2021 EDCTU]. All participants provided informed consent prior to their participation.

**Figure 1.**
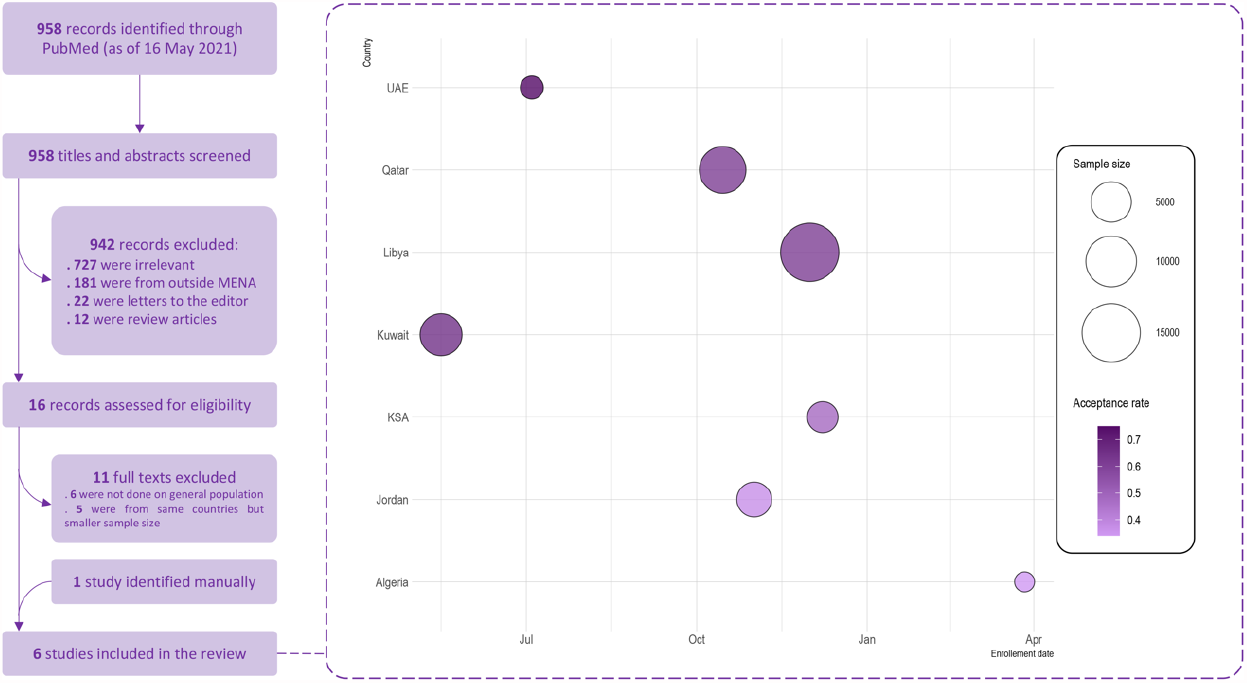
COVID-19 vaccine acceptance in the Arab countries from the MENA region - Systematic review flowchart and findings Size of the bubbles represents the sample size. The color gradient represents the acceptance rate of vaccination reported in each study; the darker the color tone the higher the acceptance rate, and vice versa. Enrollement date is the starting date of data collection.

#### I.2. Sample size and sampling technique

The sample size (N=385) was calculated using the single proportion sample size calculation formula, to detect an unknown vaccine acceptance rate (P=50%) with 95% confidence interval (95%CI), 80% statistical power and 5% margin error, among the total Algerian population. According to the WorldOMeter estimates, based on the United Nations data, the Algerian population was 44,594,368 as of May 30, 2021 (27).

A two-stage stratified snowball sampling method was used in this study. In Stage 1, Algeria was divided into four cardinal regions (strata) including North/Center, East, West and South. In stage 2, participants who were directly reached by the investigator were solicited to disseminate the questionnaire among their acquaintances until reaching a comparable number (∼N/4) of participants in each region (stratum).

#### I.3. Instrument development and validation

The questionnaire used in the present study was designed based on previously published papers related to the vaccine acceptance (28–32). It was developed in English and translated into the Arabic language by a native speaker, considering the vocabulary specificities of the Algerian population. The questionnaire comprised the following 5 sections:

##### 1) Sociodemographic data

*including participant’s age, gender, marital status, residency* region, monthly income in Algerian Dinars (AD), educational level, occupation, living mode (alone or with family), children (yes or no), and living area (rural or urban); and whether the participant has a chronic disease or lives with someone with a chronic disease.

##### 2) Health perception

including perceived health status (1 item) and perception about COVID-19 as an illness (3 items) including perceived probability of contracting COVID-19 infection, level of fear of being infected, and perceived severity of COVID-19.

##### 3) Levels of adherence to government recommendations and preventive measures against COVID-19

including 7 items, such as social distancing, hand cleaning, care seeking behavior in case of suggestive symptoms, etc. Each of the 7 items was formulated as a Likert-type agreement scale with 5 levels, including “Strongly Disagree (score=1)”, “Disagree (2)”, “Neutral (3)”, “Agree (4)”, and “Strongly agree (5)”.

##### 4) Attitudes and beliefs towards COVID-19 vaccination: including the 5 following items

“I think that SARS-CoV-2 vaccination is effective”; “In principle, I accept to get the SARS-CoV-2 vaccination”; I will to receive the SARS-CoV-2 vaccination as soon as possible whenever it is available”; “I think that the best way to avoid the complications of COVID-19 is by being vaccinated”; “I think that SARS-CoV-2 vaccination is safe”. A 5-score Likert-type agreement scale was used to encode the answers from “Strongly disagree (score=1)” to “Strongly agree (score=5)”.

##### 5) Barriers and enablers of COVID-19 vaccine acceptance

including a predefined list of potential factors that may negatively (barriers) or positively (enablers) impact the participant’s decision to receiving the COVID-19 vaccine. The list comprised 6 barriers such as concerns regarding vaccine’s side effects, conspiracy theory beliefs, etc., and 6 enablers such as vaccination enforcement policy, recommendation by physician, etc.

The questionnaire sections and items underwent face and content validity by the research team members, with the help of two public health and epidemiology experts. Further, the questionnaire was administered in a pilot sample (n =31) to assess the clarity and full understanding of questions and items. Data collected from the pilot sample was not used in the final analysis. A copy of the Arabic or English questionnaire is available upon request from the first or corresponding author.

#### I.4. Data collection procedure

The final, validated version of the questionnaire was edited as an online survey in Google Forms. An introduction was embedded in the first page of the survey consisting of the study description, an informed consent agreement, and one question related to previous COVID-19 vaccination history (eligibility criterion). The online survey link was disseminated through social media platforms including Facebook, What’s app, and Messenger. Additionally, we distributed the survey link through specific Facebook groups targeting healthcare workers and medical students, both regarding their enrollment and to enhance the snowball sampling. Data collection was anonymous and identity collecting options of Google Forms were deactivated. We followed the Strengthening the Reporting of Observational Studies in Epidemiology statement for reporting this study (33).

#### I.5. Statistical methods

##### a) Score calculation and outcome definition

Engagement score, the primary outcome, was calculated by summing up the scores of the 5 items **(Supplemental Table 1)** from efficacy, prevention from complications, safety, acceptance, and willingness subscales; high scores indicated higher levels of engagement to the vaccination. The use of an engagement score was based on the assumption that actual engagement to vaccine is a multidimensional concept depending on the participant’s perceptions and attitudes towards the vaccine safety, efficacy, prevention from complications (items 1, 4, and 5), and declared acceptance and willingness to receive it (items 2 and 3).

Adherence score (range 7—35) was calculated by summing up the scores of the 7 items **(Supplemental Table 2)** from the Adherence Level subscale; higher scores indicated higher adherence levels to recommendations and preventive measures. The variable related to adherence level was categorized into three subcategories (Low level, medium level, and high adherence level).

##### b) Statistical analysis

Categorical variables were presented as frequency and percentage, while continuous variables were presented as mean and standard deviation (SD) in the descriptive statistical analyses. Chi-square test was used to analyze the associated between categorical variables, while Mann-Whitney U test was used to analyze the distribution of numerical, non-normally distributed variables. Bivariate correlations between numerical variables used Pearson’s correlation. Multivariate binomial logistic regression was used to analyze the determinants of COVID-19 vaccine non-engagement. A p-value<0.05 was indicative for statistical significance. Statistical analysis was performed by means of IBM’s SPSS for Windows, Version 25.0 (SPSS Inc., Chicago, IL, USA).

### II. Systematic Review

#### II.1. Database search and eligibility criteria

We conducted a systematic review in compliance with the Preferred Reporting Items for Systematic Reviews and Meta-Analyses (PRISMA) reporting guidelines (34). Medline was searched through PubMed database using the following search terms: COVID-19, SARS-CoV-2, hesitancy, acceptance, vaccine, and vaccination, to retrieve related studies published from the database inception to May 16^th^, 2021. Only studies targeting the general population and reporting COVID-19 vaccination acceptance rate and studies conducted in Arab country of the MENA region were included. Review articles, editorials, case reports and case series were excluded. Additionally, the reference list of included articles was scrutinized to identify extra articles.

#### II.2. Study Selection

Two authors independently screened titles and abstracts of retrieved articles against the inclusion/exclusion criteria. Full-texts of potentially eligible articles were further assessed by two authors for final decision. Discrepancies were resolved via discussion. In the case of multiple reports from the same country, the one containing the greatest amount of information (for example, largest sample size) was included in the review.

#### II.3. Data extraction

Three investigators extracted data from relevant articles using a data extraction form. The collected data included the author’s name, study country, study period, sampling method, sample size, percentages of males and older age, acceptance rate, the predictors for COVID-19 vaccine acceptance and avoidance. A fourth experienced investigator double-checked all collected evidence for accuracy.

## Results

### Sociodemographic characteristics

A total 1,019 respondents were included, with equal distribution across the four cardinal regions in Algeria. Of these, 545 (54%) were female, 650 (64%) were aged 18-29 years, and 500 (49%) were in the healthcare sector including medical students (36%) or healthcare professionals (13%). Thus, majority were single (70%) and had high educational level (84%). Regarding comorbidities, 136 (13.3%) had a chronic disease and 531 (52.1%) were living with at least one family member having a chronic disease. Otherwise, 87.0% of the participants rated their own health status to be good or excellent (**Table 1**).

**Table 1.** Sociodemographic characteristics and answering patterns to different questionnaire scales in total population and by comparison between healthcare workers vs medical students vs the general population

| Characteristics | Total, n (%) | General Population, n (%) | Healthcare workers, n (%) | Medical students, n (%) | P-value |
| --- | --- | --- | --- | --- | --- |
| **Total** | **1019** | **519** | **136** | **364** |  |
| **Age** |  |  |  |  | **<0.001** |
| More than 60 | 54 (05%) | 52 (10%) | 2 (01%) | 0 (0%) |  |
| 40 - 59 | 107 (11%) | 99 (19%) | 7 (05%) | 1 (0.2%) |  |
| 30 - 39 | 208 (20%) | 174 (34%) | 32 (24%) | 2 (1%) |  |
| 18 - 29 | 650 (64%) | 194 (37%) | 95 (70%) | 361 99%) |  |
| **Gender** |  |  |  |  | **<0.001** |
| Males | 474 (47%) | 306 (59%) | 42 (31%) | 126 (35%) |  |
| Female | 545 (54%) | 213 (41%) | 94 (69%) | 238 (65%) |  |
| **Region** |  |  |  |  | **<0.001** |
| Center | 250 (25%) | 146 (28%) | 28 (21%) | 76 (21%) |  |
| East | 257 (25%) | 107 (21%) | 32 (24%) | 118 (32%) |  |
| Ouest | 252 (25%) | 112 (22%) | 42 (31%) | 98 (27%) |  |
| South | 260 (26%) | 154 (30%) | 34 (25%) | 72 (20%) |  |
| **Area** |  |  |  |  | **0.651** |
| Urban | 825 (81%) | 417 (80%) | 114 (84%) | 294 (81%) |  |
| Rural | 194 (19%) | 102 (20%) | 22 (16%) | 70 (19%) |  |
| **Marital status** |  |  |  |  | **<0.001** |
| Ever married | 307 (30%) | 262 (50%) | 38 (28%) | 7 (02%) |  |
| Never married | 712 (70%) | 257 (50%) | 98 (72%) | 357 (98%) |  |
| **House setting** |  |  |  |  | **0.001** |
| With family | 962 (94%) | 477 (92%) | 129 (95%) | 356 (98%) |  |
| Alone | 57 (6%) | 42 (8%) | 7 (5%) | 8 (2%) |  |
| **Income** |  |  |  |  | **<0.001** |
| > 100K DA | 199 (20%) | 95 (18%) | 33 (24%) | 71 (20%) |  |
| 50K – 100K DA | 347 (34%) | 157 (30%) | 62 (46%) | 128 (35%) |  |
| < 50K DA | 473 (46%) | 267 (51%) | 41 (30%) | 165 (45%) |  |
| **Children** |  |  |  |  | **<0.001** |
| No | 763 (75%) | 296 (57%) | 106 (78%) | 361 99%) |  |
| Yes | 256 (25%) | 223 (43%) | 30 (22%) | 3 (1%) |  |
| **Having chronic disease** |  |  |  |  | **<0.001** |
| No | 883 (87%) | 427 (82%) | 120 (88%) | 336 (92%) |  |
| Yes | 136 (13%) | 92 (18%) | 16 (12%) | 28 (08%) |  |
| **Living with someone who has a chronic disease** |  |  |  |  | **0.863** |
| No | 488 (48%) | 246 (47%) | 68 (50%) | 174 (48%) |  |
| Yes | 531 (52%) | 273 (53%) | 68 (50%) | 190 (52%) |  |
| **Perceived health status** |  |  |  |  | **0.023** |
| Below average | 131 (13%) | 439 (85%) | 126 (93%) | 323 (98%) |  |
| Good or excellent | 888 (87%) | 80 (15%) | 10 (7%) | 41 (11%) |  |
| **Fear of getting the disease** |  |  |  |  | **0.011** |
| No | 144 (14%) | 79 (15%) | 9 (07%) | 56 (15%) |  |
| Got the disease | 164 (16%) | 80 (15%) | 33 (24%) | 51 (14%) |  |
| Yes | 711 (70%) | 360 (69%) | 94 (69%) | 257 (71%) |  |
| **Perception of COVID-19 severity** |  |  |  |  | **0.013** |
| Low | 439 (43%) | 244 (47%) | 50 (37%) | 145 (40%) |  |
| Moderate | 317 (31%) | 161 (31%) | 50 (37%) | 106 (29%) |  |
| High | 263 (26%) | 114 (22%) | 36 (36%) | 113 (31%) |  |
| **Level of Adherence to preventive measures** |  |  |  |  | **0.024** |
| Low | 93 (9%) | 50 (10%) | 7 (05%) | 36 (10%) |  |
| Moderate | 491 (48%) | 245 (47%) | 56 (41%) | 190 (52%) |  |
| High | 435 (43%) | 224 (43%) | 73 (54%) | 138 (38%) |  |

### History of and perceptions towards COVID-19 infection

Majority participants (70.0%) declared fearing to contract COVID-19, and 16.0% reported a positive history of COVID-19 infection. On the other hand, only 263 (26.0%) perceived the infection to be severe, while 43.0% believed the disease had no severity. Regarding preventive measures, almost half the participants (48.0%) had moderate level of adherence, while 43.0% had high level (**Table 1**).

### Engagement towards COVID-19 vaccine

Overall, we observed low agreement levels regarding vaccine safety (21%), effectiveness (30%), and efficiency to avoid complications (32%). Likewise, a minority declared accepting COVID-19 vaccine (26%) or willing to take it (21%). Paradoxically, there were lower levels of agreement regarding vaccine safety (14% vs 25% and 26%), as well as declared acceptance (21% vs 28% and 31%) and willingness (15% vs 24% and 25%), among healthcare professionals compared with the general population and medical students respectively (p<0.001). Using the engagement score 15 (median) as cutoff, two-third of the participants had low likelihood of engagement (engagement score≤15, 66%) (**Table 2**).

**Table 2.** Engagement towards COVID-19 vaccine in total population and by comparison between healthcare workers vs medical students vs the general population

| Item / agreement level | Total, n (%) | General Population, n (%) | Healthcare workers, n (%) | Medical students, n (%) | P-value |
| --- | --- | --- | --- | --- | --- |
| **I think that SARS-CoV-2 vaccination, whenever available, would be safe** |  |  |  |  | **<0.001** |
| Strongly disagree | 193 (19%) | 113 (22%) | 14 (10%) | 66 (18%) |  |
| Disagree | 136 (13%) | 73 (14%) | 11 (08%) | 52 (14%) |  |
| Neutral | 473 (46%) | 203 (39%) | 75 (55%) | 195 (54%) |  |
| Agree | 184 (18%) | 108 (21%) | 29 (21%) | 47 (13%) |  |
| Strongly agree | 33 (3%) | 22 (04%) | 7 (05%) | 4 (01%) |  |
| **I think that SARS-CoV-2 vaccination is effective to prevent infection.** |  |  |  |  | **0.008** |
| Strongly disagree | 150 (15%) | 89 (17%) | 14 (10%) | 47 (13%) |  |
| Disagree | 167 (16%) | 95 (18%) | 19 (14%) | 53 (15%) |  |
| Neutral | 399 (39%) | 179 (34%) | 56 (41%) | 164 (45%) |  |
| Agree | 266 (26%) | 131 (25%) | 41 (30%) | 94 (26%) |  |
| Strongly agree | 37 (4%) | 25 (5%) | 6 (4%) | 6 (2%) |  |
| **I think that the best way to avoid the complications of COVID-19 is by getting vaccinated.** |  |  |  |  | **0.005** |
| Strongly disagree | 172 (17%) | 101 (19%) | 12 (09%) | 59 (16%) |  |
| Disagree | 196 (19%) | 103 (20%) | 29 (21%) | 64 (18%) |  |
| Neutral | 319 (31%) | 156 (30%) | 44 (32%) | 119 (33%) |  |
| Agree | 268 (26%) | 118 (23%) | 40 (29%) | 110 (30%) |  |
| Strongly agree | 64 (6%) | 41 (8%) | 11 (8%) | 12 (3%) |  |
| **In principle, I accept to get the SARS-CoV-2 vaccination.** |  |  |  |  | **<0.001** |
| Strongly disagree | 285 (28%) | 170 (33%) | 21 (15%) | 94 (26%) |  |
| Disagree | 190 (19%) | 78 (15%) | 32 (24%) | 80 (22%) |  |
| Neutral | 279 (27%) | 123 (24%) | 41 (30%) | 115 (32%) |  |
| Agree | 201 (20%) | 104 (20%) | 33 (24%) | 64 (18%) |  |
| Strongly agree | 64 (6%) | 44 (8%) | 9 (7%) | 11 (3%) |  |
| **I will receive the SARS-CoV-2 vaccination as soon as possible whenever it is available.** |  |  |  |  | **<0.001** |
| Strongly disagree | 326 (32%) | 181 (35%) | 25 (18%) | 120 (33%) |  |
| Disagree | 195 (19%) | 79 (15%) | 33 (24%) | 83 (23%) |  |
| Neutral | 280 (27%) | 132 (25%) | 43 (32%) | 105 (29%) |  |
| Agree | 157 (15%) | 84 (16%) | 29 (21%) | 44 (12%) |  |
| Strongly agree | 61 (6%) | 43 (8%) | 6 (4%) | 12 (3%) |  |
| **Likelihood of engagement** |  |  |  |  | **0.145** |
| High (engaged) | 342 (34%) | 181 (35%) | 52 (38%) | 109 (30%) |  |
| Low (non-engaged) | 677 (66%) | 338 (65%) | 84 (62%) | 255 (70%) |  |

### Barriers and enablers of COVID-19 vaccine acceptance

The barriers and enablers of COVID-19 vaccine acceptance are depicted in **Figure 2**. Concern about vaccine side effects was the most commonly reported barrier to COVID-19 vaccine acceptance (72.0%), followed by skepticism regarding vaccine efficacy in preventing the infection (29.0%) and beliefs in the conspiracy theory (23.4%). Regarding enablers, exigence for more efficacy and safety studies was the most commonly reported (48.3%), followed by condition that the vaccine be recommended by the physician (16.3%) or become mandatory (12.9%).

**Figure 2.**
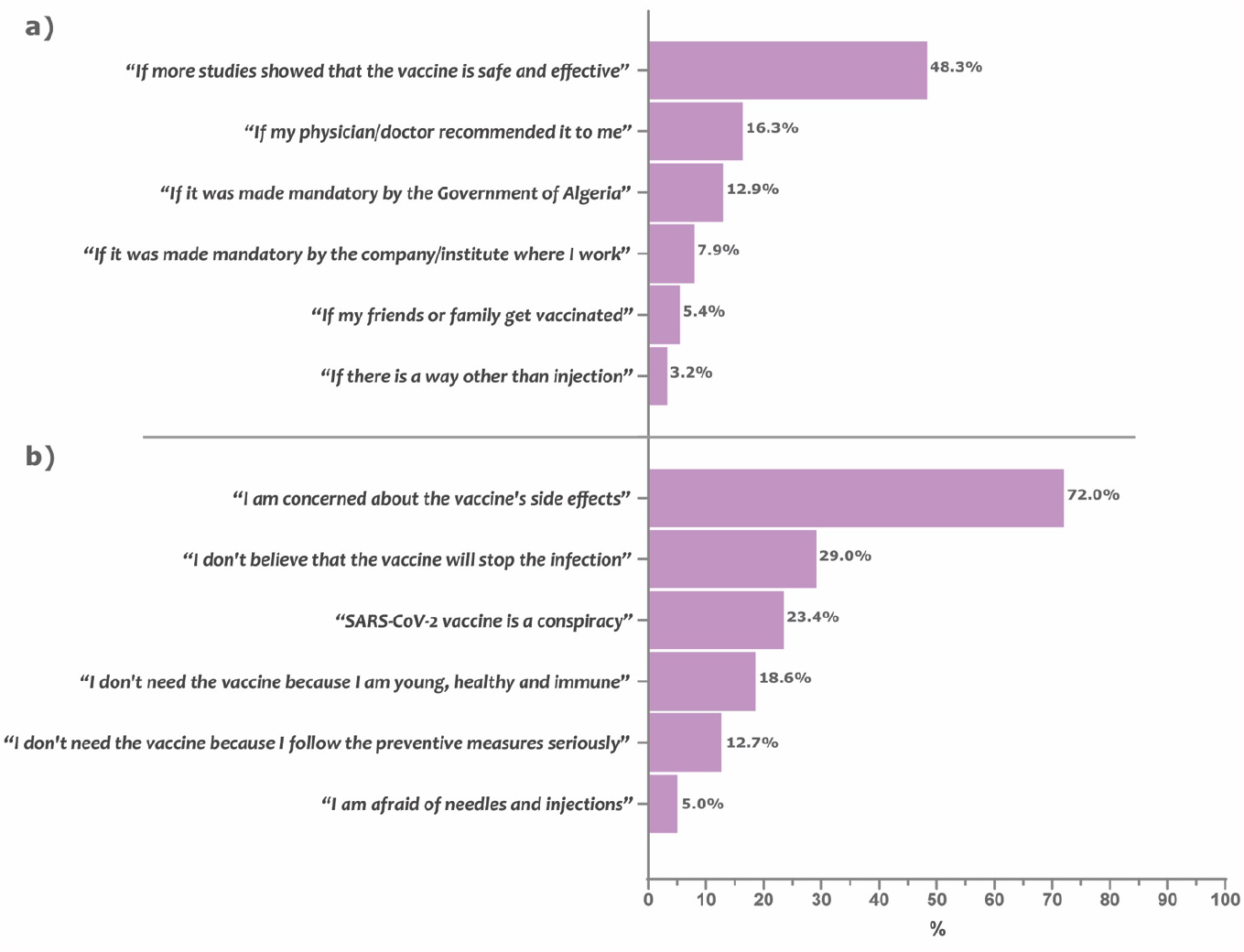
Enablers and barriers of COVID-19 vaccine acceptance Bars represent the percentage of participants who reported the given item as being a determining enabler (a) or barrier (b) for acceptance of the COVID-19 vaccine uptake.

### Factors associated with COVID-19 vaccine nonengagement

In unadjusted models, younger age, female gender, unmarried status, higher income, and highly perceived healthiness were associated with higher likelihood for nonengagement to vaccine (OR>1, p<0.05), by reference to their respective counterparts. On the other hand, having children, being afflicted with a chronic disease, highly perceived severity of COVID-19 and fear from being infected were associated with lower likelihood for nonengagement to vaccine (OR<1, p<0.05), by reference to their respective counterparts. Further, the level of adherence to preventive measure was inversely associated with nonengagement to vaccine (**Table 3**).

**Table 3.** Factors associated with vaccine engagement levels

| Parameter / category | Total<br>(n = 1019) | Engagement<br>score | Non-engagement (Engagement score≤15) |  |  |  |  |
| --- | --- | --- | --- | --- | --- | --- | --- |
|  | N (%) | Mean ± SD | Rate, N (%) | Unadjusted OR<br>(95%CI) | P-value | Adjusted OR<br>(95%CI) | P-value |
| **Age** |  |  |  |  |  |  |  |
| More than 60 y | 54 (5.3%) | 15.91 ± 6.77 | 23 (42.6%) | ref |  | ref |  |
| 40 – 59 y | 107 (10.5%) | 13.41 ± 5.96 | 66 (61.7%) | **2.17 (1.12 - 4.22)** | **0.023** | 1.77 (0.82 - 3.83) | 0.145 |
| 30 – 39 y | 208 (20.4%) | 13.13 ± 6.08 | 134 (64.4%) | **2.44 (1.33 - 4.49)** | **0.004** | 1.46 (0.68 - 3.13) | 0.329 |
| 18 – 29 y | 650 (63.8%) | 13.41 ± 4.57 | 454 (69.8%) | **3.12 (1.78 - 5.49)** | **< 0.001** | 1.39 (0.61 - 3.17) | 0.432 |
| **Gender** |  |  |  |  |  |  |  |
| Males | 474 (46.5%) | 13.90 ± 5.74 | 284 (59.9%) | ref |  | ref |  |
| Female | 545 (53.5%) | 13.13 ± 4.70 | 393 (72.1%) | **1.73 (1.33 - 2.25)** | **< 0.001** | **2.31 (1.68 - 3.18)** | **< 0.001** |
| **Region** |  |  |  |  |  |  |  |
| Center | 250 (24.5%) | 13.27 ± 5.80 | 163 (65.2%) | ref |  | - |  |
| East | 257 (25.2%) | 14.14 ± 5.06 | 158 (61.5%) | 0.85 (0.59 - 1.22) | 0.385 |  |  |
| Ouest | 252 (24.7%) | 13.40 ± 4.99 | 173 (68.7%) | 1.17 (0.81 - 1.70) | 0.411 |  |  |
| South | 260 (25.5%) | 13.13 ± 4.98 | 183 (70.4%) | 1.27 (0.87 - 1.84) | 0.211 |  |  |
| **Area** |  |  |  |  |  |  |  |
| Urban | 825 (81%) | 13.60 ± 5.19 | 542 (65.7%) | ref |  | - |  |
| Rural | 194 (19%) | 12.98 ± 5.36 | 135 (69.6%) | 1.20 (0.85 - 1.68) | 0.302 |  |  |
| **Marital status** |  |  |  |  |  |  |  |
| Ever married | 307 (30.1%) | 13.92 ± 5.92 | 184 (59.9%) | ref |  | ref |  |
| Never married | 712 (69.9%) | 13.30 ± 4.88 | 493 (69.2%) | **1.51 (1.14 - 1.99)** | **0.004** | 1.10 (0.59 - 2.04) | 0.76 |
| **Level of education** |  |  |  |  |  |  |  |
| Low level | 56 (5.5%) | 12.20 ± 6.45 | 37 (66.1%) | ref |  | - |  |
| Medium level | 110 (10.8%) | 13.27 ± 5.88 | 70 (63.6%) | 0.89 (0.46 - 1.77) | 0.757 |  |  |
| High level | 853 (83.7%) | 13.60 ± 5.03 | 570 (66.8%) | 1.03 (0.58 - 1.83) | 0.908 |  |  |
| **House setting** |  |  |  |  |  |  |  |
| With family | 962 (94.4%) | 13.53 ± 5.17 | 642 (66.7%) | ref |  | - |  |
| Alone | 57 (5.6%) | 12.68 ± 6.04 | 35 (61.4%) | 0.79 (0.46 - 1.37) | 0.408 |  |  |
| **Living with someone who has a chronic disease** |  |  |  |  |  |  |  |
| No | 488 (47.9%) | 13.26 ± 5.17 | 334 (68.4%) | ref |  | - |  |
| Yes | 531 (52.1%) | 13.70 ± 5.26 | 343 (64.6%) | 0.84 (0.65 - 1.09) | 0.194 |  |  |
| **Having chronic disease** |  |  |  |  |  |  |  |
| No | 883 (86.7%) | 13.34 ± 5.15 | 598 (67.7%) | ref |  | ref |  |
| Yes | 136 (13.3%) | 14.45 ± 5.57 | 79 (58.1%) | **0.66 (0.46 - 0.96)** | **0.027** | 0.88 (0.56 - 1.38) | 0.579 |
| **Job** |  |  |  |  |  |  |  |
| Unemployed | 144 (14.1%) | 12.76 ± 5.86 | 100 (69.4%) | ref |  | ref |  |
| Healthcare sector | 136 (13.3%) | 14.66 ± 4.61 | 84 (61.8%) | 0.71 (0.43 - 1.17) | 0.177 | 0.60 (0.32 - 1.02) | 0.057 |
| Public sector | 165 (16.2%) | 13.60 ± 5.57 | 106 (64.2%) | 0.79 (0.49 - 1.27) | 0.334 | 0.80 (0.46 - 1.40) | 0.438 |
| Privat sector | 122 (12%) | 13.84 ± 6.24 | 70 (57.4%) | **0.59 (0.36 - 0.98)** | **0.042** | **0.53 (0.29 - 0.97)** | **0.038** |
| Student | 364 (35.7%) | 13.20 ± 4.61 | 255 (70.1%) | 1.03 (0.67 - 1.57) | 0.892 | 0.67 (0.39 - 1.15) | 0.147 |
| Others | 88 (8.6%) | 13.34 ± 4.99 | 62 (70.5%) | 1.05 (0.59 - 1.87) | 0.871 | 1.07 (0.55 - 2.06) | 0.851 |
| **Income** |  |  |  |  |  |  |  |
| > 100K AD | 199 (19.5%) | 14.69 ± 5.22 | 117 (58.8%) | ref |  | ref |  |
| 50K – 100K AD | 347 (34.1%) | 13.48 ± 5.02 | 235 (67.7%) | **1.47 (1.03 - 2.11)** | **0.036** | 1.47 (0.99 - 2.17) | 0.051 |
| < 50K AD | 473 (46.4%) | 12.98 ± 5.29 | 325 (68.7%) | **1.54 (1.09 - 2.17)** | **0.014** | 1.34 (0.92 - 1.95) | 0.132 |
| **Children** |  |  |  |  |  |  |  |
| No | 763 (74.9%) | 13.30 ± 4.96 | 527 (69.1%) | ref |  | ref |  |
| Yes | 256 (25.1%) | 14.04 ± 5.92 | 150 (58.6%) | **0.63 (0.47 - 0.85)** | **0.002** | 0.73 (0.40 - 1.35) | 0.315 |
| **Fear of getting the disease** |  |  |  |  |  |  |  |
| No | 144 (14.1%) | 11.12 ± 5.37 | 116 (80.6%) | ref |  | ref |  |
| Got the disease | 164 (16.1%) | 13.70 ± 4.58 | 114 (69.5%) | **0.55 (0.32 - 0.94)** | **0.027** | 0.68 (0.38 - 1.21) | 0.19 |
| Yes | 711 (69.8%) | 13.92 ± 5.21 | 447 (62.9%) | **0.41 (0.26 - 0.63)** | **< 0.001** | **0.56 (0.35 - 0.91)** | **0.018** |
| **Perception of COVID-19 severity** |  |  |  |  |  |  |  |
| Null | 439 (43.1%) | 12.36 ± 5.50 | 318 (72.4%) | ref |  | ref |  |
| Medium | 317 (31.1%) | 14.38 ± 4.79 | 194 (61.2%) | **0.60 (0.44 - 0.82)** | **0.001** | 0.76 (0.52 - 1.09) | 0.134 |
| High | 263 (25.8%) | 14.30 ± 4.90 | 165 (62.7%) | **0.64 (0.46 - 0.89)** | **0.007** | **0.66 (0.47 - 0.92)** | **0.014** |
| **Health perception** |  |  |  |  |  |  |  |
| Below average | 131 (12.9%) | 14.17 ± 5.67 | 76 (58.0%) | ref |  | ref |  |
| Good/excellent | 888 (87.1%) | 13.39 ± 5.15 | 601 (67.7%) | **1.52 (1.04 - 2.20)** | **0.03** | 1.45 (0.94 - 2.24) | 0.097 |
| **Level of Adherence to preventive measures** |  |  |  |  |  |  |  |
| High level | 435 (42.7%) | 15.06 ± 5.24 | 243 (55.9%) | ref |  | ref |  |
| Medium level | 491 (48.2%) | 12.78 ± 4.79 | 352 (71.7%) | **2.00 (1.52 - 2.63)** | **< 0.001** | **2.07 (1.54 - 2.78)** | **< 0.001** |
| Low adherence | 93 (9.1%) | 9.86 ± 4.74 | 82 (88.2%) | **5.89 (3.05 - 11.36)** | **< 0.001** | **6.93 (3.46 - 13.87)** | **< 0.001** |
| 95% CI. 95% confidence interval; AD. Algerian dinar; N. number; OR. odds ratio; SD. standard deviation; y. years; *. adjusted for age. gender. marital status. having chronic disease. job. income. having children. fear from getting the disease. perception of severity of the disease. health perception and level of adherence to preventive measures. |  |  |  |  |  |  |  |

The Adjusted model showed that the likelihood for nonengagement was independently associated with female gender (OR=2.31; 95%CI: 1.68 - 3.18, p<0.001), medium (OR=2.07, 95%CI: 1.54 - 2.78, p< 0.001) and low adherence level to preventive measures (OR=6.93; 95%CI: 3.46 - 13.87, p< 0.001), work in private sector (OR=0.53; 95%CI: 0.29 - 0.97, p=0.038), high perceived COVID-19 severity (OR=0.66; 95%CI: 0.47 - 0.92, p=0.014), and fear from contracting the disease (OR=0.56; 95%CI: 0.35 - 0.91, p=0.018) (**Table 3, Figure 3**).

**Figure 3.**
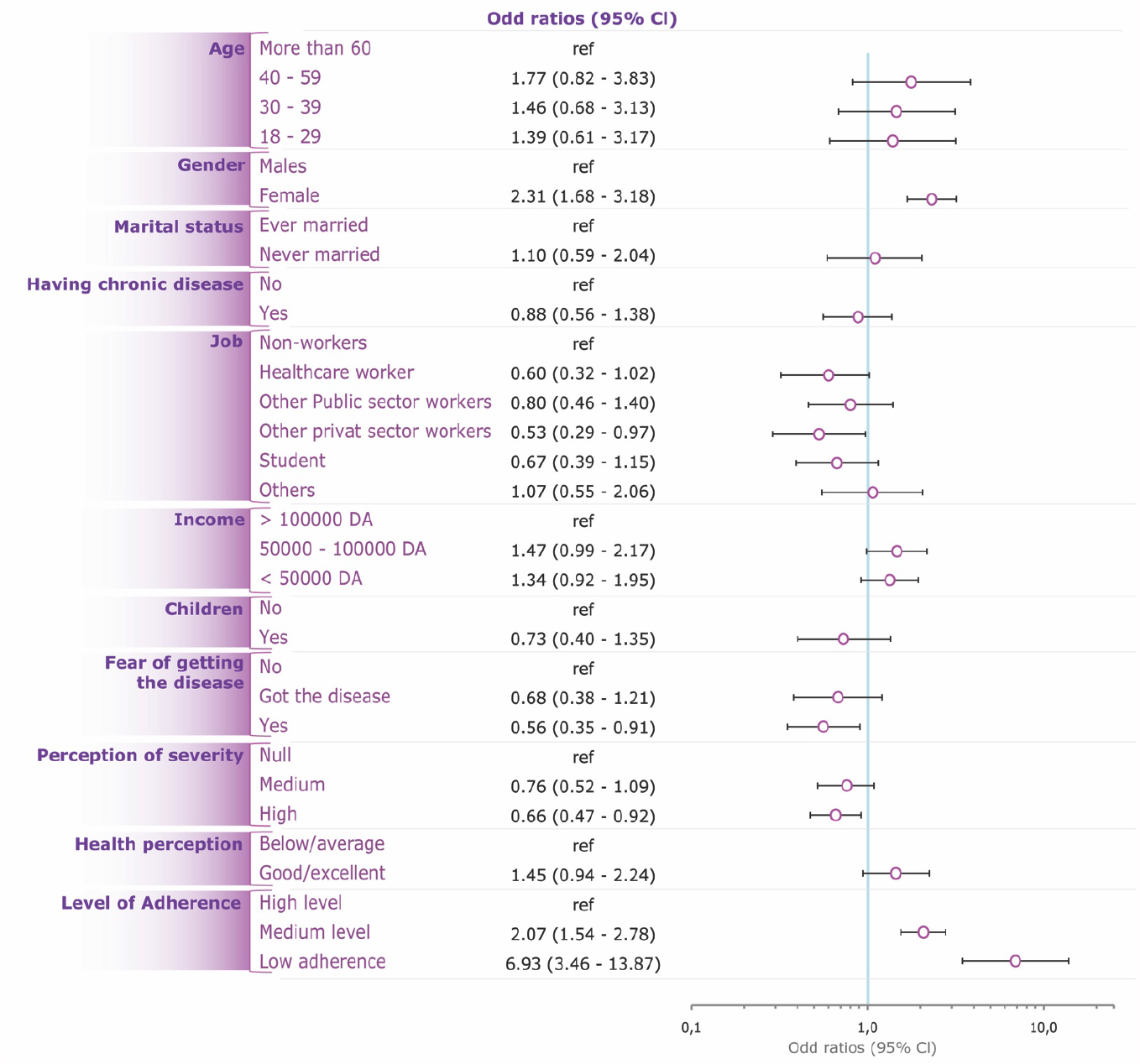
Predictors of nonengagement to COVID-19 vaccine

### COVID-19 acceptance in Arab countries from MENA region – Results of the systematic review

A total of 6 studies were included in this systematic review, including 1,019-15,087 participants. Eleven studies were excluded, out of which 6 were not conducted among general population, and 5 studies comprised a smaller sample size as shown in **Figure 1**. The included studies were conducted in United Arab Emirates (UAE), Kuwait, Qatar, Libya, Kingdom of Saudi Arabia (KSA), and Jordan (**Table 4**). All studies were internet-based, nationwide surveys; 3 studies (28– 30) were conducted only amongst the general population, while the remaining comprised general population and healthcare workers (31,32,35). The highest COVID-19 vaccine acceptance rate (75%) was reported in UAE (28), followed by Kuwait (65%) (29), Qatar (61%) (31), and Libya (61%) (32). Predictors of vaccine acceptance varied between the studies, and included adherence to government recommendations, married status, positive COVID-19 status, having friends died or infected with COVID-19, high income, fear of contracting COVID-19, perception of high severity, and private sector workers. History of flu vaccination was a positive predictor of COVID-19 vaccination in 3 studies by Alabdulla et al. (31), Alfageeh et al. (30), and El-Elimat et al. (35). Female gender was a significant predictor for vaccine avoidance in the study by Alfageeh et al. (30). Other vaccine avoidance predictors that were reported comprised younger age, self-employment, safety concerns, conspiracy theory, and low and medium adherence to COVID-19 preventive measures.

**Table 4.**
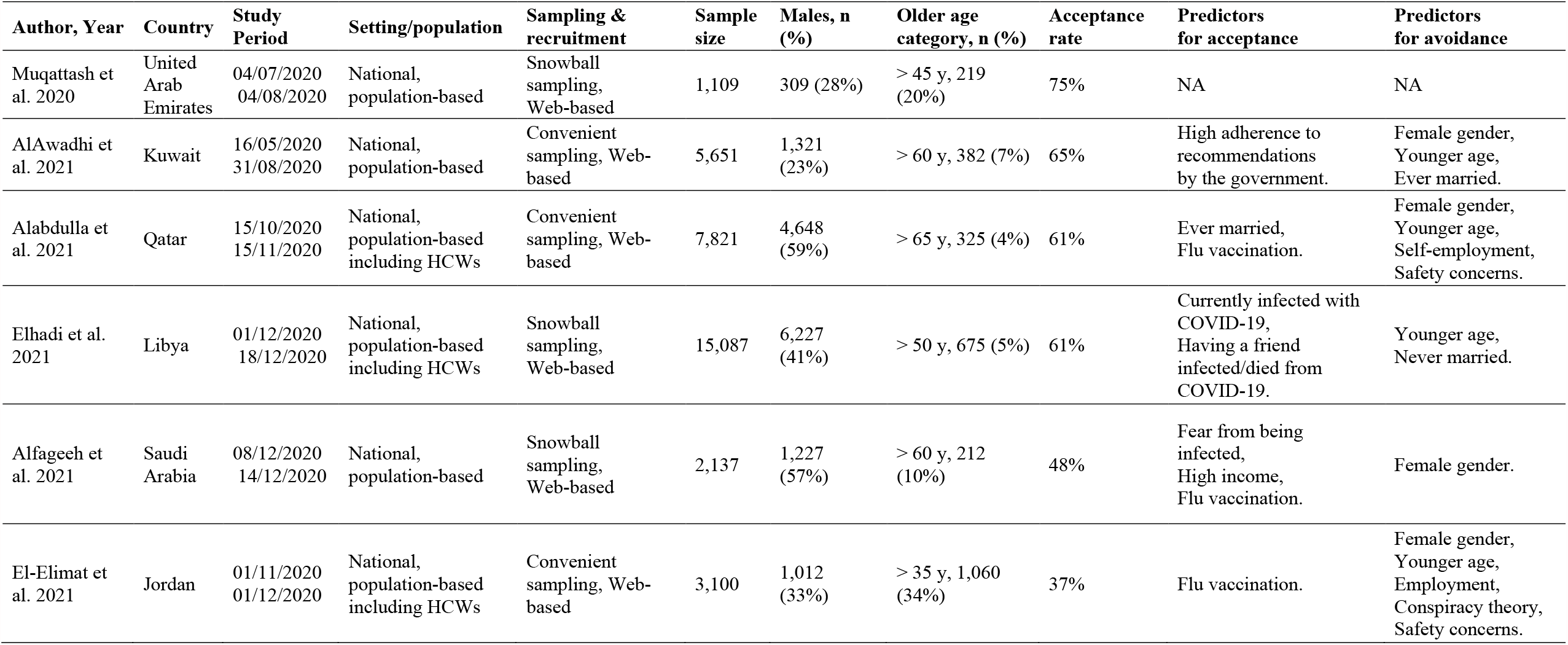
Characteristics of studies included from the MENA regions

## Discussion

This is the first nationwide study addressing the Algerian population’s attitude toward the SARS-CoV-2 vaccine. Using a multidimensional model to measure the likelihood of engagement to vaccination, our study revealed that only 34% of the participants would be engaged to receive the COVID-19 vaccines. The Adjusted regression analysis demonstrated multiple predictors for non-engagement, including female gender, and low/intermediate levels of adherence to preventive measures, whereas a high perception of the disease severity and fear of being infected predicted vaccine acceptance. Additionally, the systematic review findings suggested that Algeria had the lowest vaccine acceptance rate in comparison with other MENA countries, where acceptance rates ranged from 37.4% in Jordan (35) and 75% in the UAE (28). In comparison with Europe, the lowest acceptance rate of 53.7%, reported in Italy (36), was relatively higher than the acceptance rate observed in our study.

High perceived severity of COVID-19 was among the independent risk factors for engagement; however, only 25.8% of participants perceived the disease to be severe. Regardless of the acceptability of the vaccine, the severity of the disease will affect the vaccination intention. Perception about the disease severity may be assimilated to a personal opinion or belief regarding the level of hazard or exposure to the crisis and the extent of its adverse impact on the individual (37). In the case of COVID-19, but not specifically, the risk perception may change over time and is further determined by the individual’s awareness about and interpretation of the relationship between the virus/pandemic and the observed undesirable effects – and such interpretation may be biased or distorted by other opinions, (mis)beliefs and (mis)conceptions. A theoretical approach by Cori et al. (38), suggested that both risk perception and fear of COVID-19 are determined by cognitive factors, and the author mentioned four key factors including knowledge about the disease/virus, visibility of the risk, trust in the authorities and healthcare institutions, and voluntary exposure to the virus/infection. The aforementioned factors may be modified by means of awareness raising campaigns and authoritarian corrective or restrictive measures, aiming at enhancing the risk perception and ultimately increasing the vaccination rates. Evidence from previous data suggests that risk perception about COVID-19 increased in the lockdown phase and decreased in the re-opening phases (39), which was positively associated with the change in vaccine acceptance rate. At the time when the present study was conducted, the country was in a post-re-opening phase, which may explain the low engagement rates observed. Another longitudinal study from the US assessed the trend of people’s attitude towards vaccine, between March and August 2020, and showed heterogenous results with perceived severity of the disease being one of the determinants of the vaccine acceptance. Furthermore, authors demonstrated that the trends in both risk perception and vaccine acceptance were likely to be determined by the individual’s specific political positions and exposure to media (40). Such observation supports the importance of correcting the cognitive and behavioral factors at the population’s level to enhance the vaccine uptake.

Similar to other reports from the MENA region, including Kuwait (29), Qatar (31), KSA (30), and Jordan (35), men were more likely to accept the Covid-19 vaccine in Algeria. This can be explained by the increased severity of the disease among men and the higher mortality reported in majority countries (41,42). This statement was extensively mediatized and may have played a role in men’s motive to vaccination, developing a relatively more positive attitude towards vaccine. While such an explanation requires further evidence, notably the associated levels of awareness about the specific health risks on males, other factors may explain the less negative attitude among males that was found in the present study. Among these factors are the impact of the pandemic and restrictive measures on incomes and businesses, which may be more perceived by males in some societies. This explanation may be in line with the significant association of vaccine engagement with being married and having children that were found in the unadjusted analysis in the present study.

However, past research data showed conflicting results about gender. A global survey including 13,426 individuals in 19 countries with a high COVID-19 burden showed that men were relatively less likely to have a positive attitude towards vaccination than women (43). Another study showed that women in Russia and Germany had higher acceptance rates of the SARS-CoV-2 vaccines than men (44). This phenomenon has been named “*the Covid-19 gender paradox*” (45). This gender difference can be explained by multidimensional psychological, social, cultural, and environmental influences. Further research may be required to determine the gender-specific factors associated with acceptance or refusal of the vaccine, which would enable designing targeted awareness campaigns with gender-specific messages to enhance the vaccine acceptance rates in both genders.

There is a remarkable similarity between the engagement rates of the general population (35%) and healthcare workers (38%) in the present study, which is an issue of big concern as it may constitute a significant barrier to the national vaccine campaign. Indeed, the practitioner’s vaccine hesitancy influences the vaccination attitudes of the patients (46). When providers are unsure of the safety of the vaccine, they are unable to recommend it to the general population. Such an issue should be considered at the critical level by the health authorities, and corrective measures are warranted urgently to increase awareness among health providers. Furthermore, this study showed comparable patterns of safety concerns about the vaccine in the two subgroups, i.e. health workers versus the general population (75% and 73%, respectively). This indicates the consistency of the popular misconceptions about the COVID-19 vaccine across all categories of the studied population and highlights the need for a comprehensive awareness-raising campaign at the national scale.

Other notable factors of vaccine refusal include fear of the side effects and concern about the efficiency of the vaccines. Similar concerns have been reported in other countries such as Jordan (35,47) and the USA (47). Arguably, these concerns may be comprehensible, considering the rapid vaccine development process, the novelty of the mRNA technology used in some vaccines, and the public mediatization of the vaccine side effects; all exposing the population to massive misinformation notably in the social media (48,49). This could be related to the decreasing acceptance rate over time in the MENA region as shown in the systematic review part of the study. Hence there is a crucial need to implement effective strategies to correct the popular misconceptions regarding the vaccine’s safety.

This study also highlighted the positive association between the level of adherence to preventive measures and vaccine acceptability. This observation is in accordance with another MENA region study in Kuwait (29), reporting that high adherence to the governmental recommendations was an important predictor for vaccine uptake. Both low adherence to preventive measures and adverse attitude towards vaccines could reflect adherence to the conspiracy theory, and this was observed among 23.4% of the avoidant group. Conspiracy theories have been associated with vaccine hesitancy as a result of mistrust between the public and the government policymakers (50).

## Strengths and limitations

One of the strengths of the present study is the use of a multidimensional model to define vaccine engagement based on a conceptual framework combining perceived vaccine effectiveness and safety with self-declared acceptance and willingness. This combination is assumed to be more reliable than using self-declared acceptance and willingness, as perceived safety and efficacy are less subject to social desirability bias. Yet, the scale requires further validation to support this assumption. On the other hand, there are no validated instruments to assess attitudes toward the COVID-19 vaccines, and the relevant studies principally used various formulations of self-declared willingness or preparedness, which is limited by the high risk of negative or positive social desirability bias. Future research is recommended in this regard to design a validated scale to measure vaccine acceptance based on a strong model, which will enhance the quality and comparability of the findings. Another strong point of this study is that participants were equally distributed from the 4 regions of the country, which supports the generalizability of the findings. Further, determinants of vaccine acceptance and avoidance were highlighted for the first time nationwide. Therefore, findings of this study can have a high impact on health authorities’ decisions for the management of vaccination campaign.

The major limitation of this study is the recruitment method of the participants, which was restricted to those who have access to the internet and an electronic device, since the questionnaire was shared online. This probably led to a selection bias, occulting a non-negligible section of the population that may have distinct characteristics. One of these characteristics is the source of information regarding COVID-19 disease and vaccine, which may be radically different in the subpopulation of internet nonusers by reference to internet users. This may result in discrepant opinions and attitudes towards the vaccine by reference to the study population. Unfortunately, no data was collected about sources of information about the vaccines, which would provide an indication about the aforementioned issue. Nevertheless, a study showed that individuals who get information from the internet are less incline to accept the COVID-19 vaccine than those who get information from healthcare workers (51).

## Conclusion

Two-third of Algerians are likely to be nonengaged for COVID-19 vaccine uptake, making them one of the least accepting public for the voluntary vaccination in the MENA region. This probably provides an explanation for the slow ascension of the vaccination curve, which constitutes a great public health concern. These findings and their interpretation should be taken into consideration by the policymakers to acknowledge and address the adverse attitude about the vaccine, notably among healthcare providers who are the vectors and major contributors of a successful vaccine policy. Vaccine awareness campaigns should be implemented to address the multiple misconceptions and enhance the levels of knowledge and perception both about the disease and the vaccine, by prioritizing target populations and engaging both healthcare workers and the general population.

## Data Availability

The data can be shared upon request from the corresponding author

---

**Supplemental Table 1:** Inter-correlation matrix of the variables used in the calculation of engagement score

|  | Item 1 | Item 2 | Item 3 | Item 4 | Item 5 |
| --- | --- | --- | --- | --- | --- |
| Item 1 | 1.00 | 0.70 | 0.68 | 0.79 | 0.80 |
| Item 2 |  | 1.00 | 0.89 | 0.75 | 0.77 |
| Item 3 |  |  | 1.00 | 0.72 | 0.74 |
| Item 4 |  |  |  | 1.00 | 0.78 |
| Item 5 |  |  |  |  | 1.00 |
| **Item 1:** I think that SARS-CoV-2 vaccination, whenever available, would be effective. |  |  |  |  |  |
| **Item 2:** In principle, I accept to get the SARS-CoV-2 vaccination. |  |  |  |  |  |
| **Item 3:** I will receive the SARS-CoV-2 vaccination as soon as possible whenever it is available. |  |  |  |  |  |
| **Item 4:** I think that the best way to avoid the complications of COVID-19 is by getting vaccinated. |  |  |  |  |  |
| **Item 5:** I think that SARS-CoV-2 vaccination, whenever available, would be safe. |  |  |  |  |  |
| Cronbach Alpha = 0.94. |  |  |  |  |  |

**Supplemental Table 2:** Inter-correlation matrix of the variables used in the calculation of adherence score

|  | Item 1 | Item 2 | Item 3 | Item 4 | Item 5 | Item 6 | Item 7 |
| --- | --- | --- | --- | --- | --- | --- | --- |
| Item 1 | 1.00 | 0.43 | 0.55 | 0.34 | 0.51 | 0.47 | 0.38 |
| Item 2 |  | 1.00 | 0.37 | 0.58 | 0.32 | 0.39 | 0.33 |
| Item 3 |  |  | 1.00 | 0.32 | 0.59 | 0.46 | 0.37 |
| Item 4 |  |  |  | 1.00 | 0.35 | 0.43 | 0.42 |
| Item 5 |  |  |  |  | 1.00 | 0.55 | 0.40 |
| Item 6 |  |  |  |  |  | 1.00 | 0.54 |
| Item 7 |  |  |  |  |  |  | 1.00 |
| **Item 1:** I don’t touch my nose, ears or mouth. |  |  |  |  |  |  |  |
| **Item 2:** I clean my hands using soap and water, or alcohol-based hand rub. |  |  |  |  |  |  |  |
| **Item 3:** I avoid shaking hands when I am outside. |  |  |  |  |  |  |  |
| **Item 4:** I cover my nose and mouth with my bent elbow or a tissue when I cough or sneeze. |  |  |  |  |  |  |  |
| **Item 5:** I maintain safe distance from anyone who is coughing or sneezing. |  |  |  |  |  |  |  |
| **Item 6:** I stay at home when I feel unwell. |  |  |  |  |  |  |  |
| **Item 7:** I seek medical attention when I have fever, cough and difficulty breathing. |  |  |  |  |  |  |  |
| Cronbach Alpha = 0.84. |  |  |  |  |  |  |  |

